# Outcomes after radioiodine treatment for thyrotoxicosis

**DOI:** 10.1101/2022.01.17.22269444

**Authors:** Mark J Bolland, Michael S Croxson

## Abstract

**Background:** Radioiodine is commonly prescribed as a permanent treatment for thyrotoxicosis. At ADHB, Auckland, New Zealand, radioiodine dose is individualised by the prescribing physician according to patient characteristics.

**Aims:** We investigated the outcomes of this approach.

**Methods:** We identified all patients receiving radioiodine for thyrotoxicosis at ADHB in 2015 and retrieved relevant clinical details.

**Results:** 222 patients were prescribed radioiodine: 147 (66%) for Graves’ disease, 58 (26%) for toxic nodular goitre, and 17 (8%) for solitary toxic nodule. For Graves’ disease, 80% had one radioiodine dose (first dose median 550 MBq, range 200-1000 MBq; total dose 200-2400 MBq), 92% had the thyrotoxicosis cured, and 83% required thyroxine post-radioiodine. For toxic nodular goitre, 93% had one dose (median 550 MBq, range 400-1000 MBq, total dose 400-1800 MBq), 93% were cured and 22% required thyroxine. For solitary toxic nodule, all had one dose (median 550 MBq, range 500-550 MBq), all were cured and 35% required thyroxine. In 69/222 (31%) patients (35% of individuals with Graves’ disease, 17% with toxic nodular goitre, and 47% with solitary toxic nodule), the most recent TSH (mean 3.2 years post-radioiodine) was elevated (30% TSH >10 mu/L, 70% TSH 4-10 mu/L).

**Conclusions:** Following radioiodine treatment, >90% of individuals have the thyrotoxicosis cured, but hypothyroidism is usual in Graves’ disease and occurs in 22-35% in toxic nodular goitre or solitary toxic nodule. Many individuals taking thyroxine after radioiodine have suboptimally controlled hypothyroidism.

## Introduction

Thyrotoxicosis is one of the most common endocrine conditions. It is most often due to autoimmune hyperthyroidism (Graves’ disease), but toxic nodular goitre or a solitary toxic nodule accounts for a substantial minority of cases, especially in older people. Antithyroid medication is the first-line treatment but does not offer a permanent cure. Radioiodine or thyroidectomy are the options available for patients seeking permanent treatment.

Surveys of endocrinologists’ management practices for Graves’ disease have been carried out in Australia and New Zealand.^1,2^ In both surveys, the use of radioiodine as a primary treatment for the first episode of Graves’ disease in a 43 year old woman was uncommon (19% and 5% respectively). In the Australian survey, participants were also asked about treatment if the patient was 71 years old, or if there had been recurrent episodes. In both situations, radioiodine was the most common treatment choice.^1^ Radioiodine practice was similar in both surveys: about 50% used a fixed dose (Australia 80% used dose of 555 MBq, New Zealand not reported), 20-34% used individualised dosing, and 40% aimed for euthyroidism after treatment.^1,2^

Despite the common occurrence of thyrotoxicosis, there are surprisingly few published data from Australia and New Zealand about the outcomes of radioiodine treatment, and in particular, whether the target of euthyroidism is realistic. From a PubMed literature search, we identified reports of rates of cure and euthyroidism following radioiodine from Newcastle,^3^ Brisbane,^4^ and Hamilton, New Zealand.^5,6^ All the studies used fixed doses (555MBq, 450-500 MBq, and 555 MBq, respectively), two reported outcomes after one dose only,^3,5^ and two reported on causes of thyrotoxicosis other than Graves’ disease.^3,5^

The Auckland District Health Board (ADHB) radioiodine clinic serves a catchment population of approximately 2 million. Prescribing of radioiodine is individualised according to patient characteristics by the prescribing physician, rather than using a fixed dose as in the four identified reports of outcome data.^3-6^ We audited the treatment of patients receiving radioiodine at ADHB to describe the outcomes and see how they compare to physician expectations of post-radioiodine euthyroidism. A secondary consequence is that the outcomes could be compared and contrasted with different approaches, such as using fixed dose radioiodine.

## Methods

All patient appointments for outpatient radioiodine at ADHB in 2015 were identified. Of 305 appointments, 4 were duplicates, 6 rescheduled or did not attend, and 7 declined or were not given radioiodine, leaving 288 potentially eligible patients for inclusion. Clinical details were retrieved from the medical records, including demographics and ethnicity, diagnosis listed by the referring clinician, laboratory tests including thyroid function tests and TSH receptor antibodies (TSH-R ab) in patients with Graves’ disease, medication usage, radioiodine dose(s), and whether thyroxine was required after radioiodine. In patients who received a second or third dose of radioiodine in 2015 or were prescribed subsequent doses after an initial dose in 2015, their records for all doses were obtained. Cure of thyrotoxicosis was defined as either development of hypothyroidism with long-term thyroxine use required, or persistent normal or elevated TSH in the absence of antithyroid medication at least 1 year after radioiodine dose. TSH was measured using different assays, but for analysis purposes, the normal range was considered to be 0.4-4.0 mU/L, and TSH was considered undetectable when the result was listed as “<value”.

At ADHB, neither thyroid scintigraphy nor isotope uptake scans to calculate doses are routinely performed prior to radioiodine treatment. Prescribing of radioiodine was individualised according to patient characteristics by the prescribing physician. Generally, dose was influenced by the size of the thyroid gland, as determined by the prescribing physician. Very high levels of TSH-R ab were also a consideration for a higher dose.

Descriptive characteristics are presented: analyses of continuous variables using mean (SD) or median (range) as appropriate, and categorical variables using frequencies. Data are presented by diagnosis and by first dose of radioiodine received. We assessed factors that might influence the development of hypothyroidism after radioiodine (pre-radioiodine medication dose, pre-radioiodine TSH, and total dose of radioiodine) using chi-square tests. All analyses were performed with R software packages (R 3.5.1, 2019, R Foundation for Statistical Computing, Vienna, Austria). This is an audit as defined by the New Zealand National Ethics Advisory Committee guidelines and therefore it did not require ethical approval.^7^ As such, ethical approval was waived by the Health and Disability Ethics Committees (HDEC) of New Zealand.

## Results

288 patients were prescribed radioiodine. The indication was cancer treatment for 58 (20%), euthyroid goitre for 7 (2%) and 1 patient was from another city with few details available. Therefore, 222 patients with available clinical records were prescribed radioiodine for thyrotoxicosis: 147 (66%) Graves’ disease, 58 (26%) toxic nodular goitre, and 17 (8%) solitary toxic nodule. Their characteristics are described in Table 1. The majority were female, and those with Graves’ disease were younger than those with toxic nodular goitre or solitary toxic nodule.

**Table 1:**
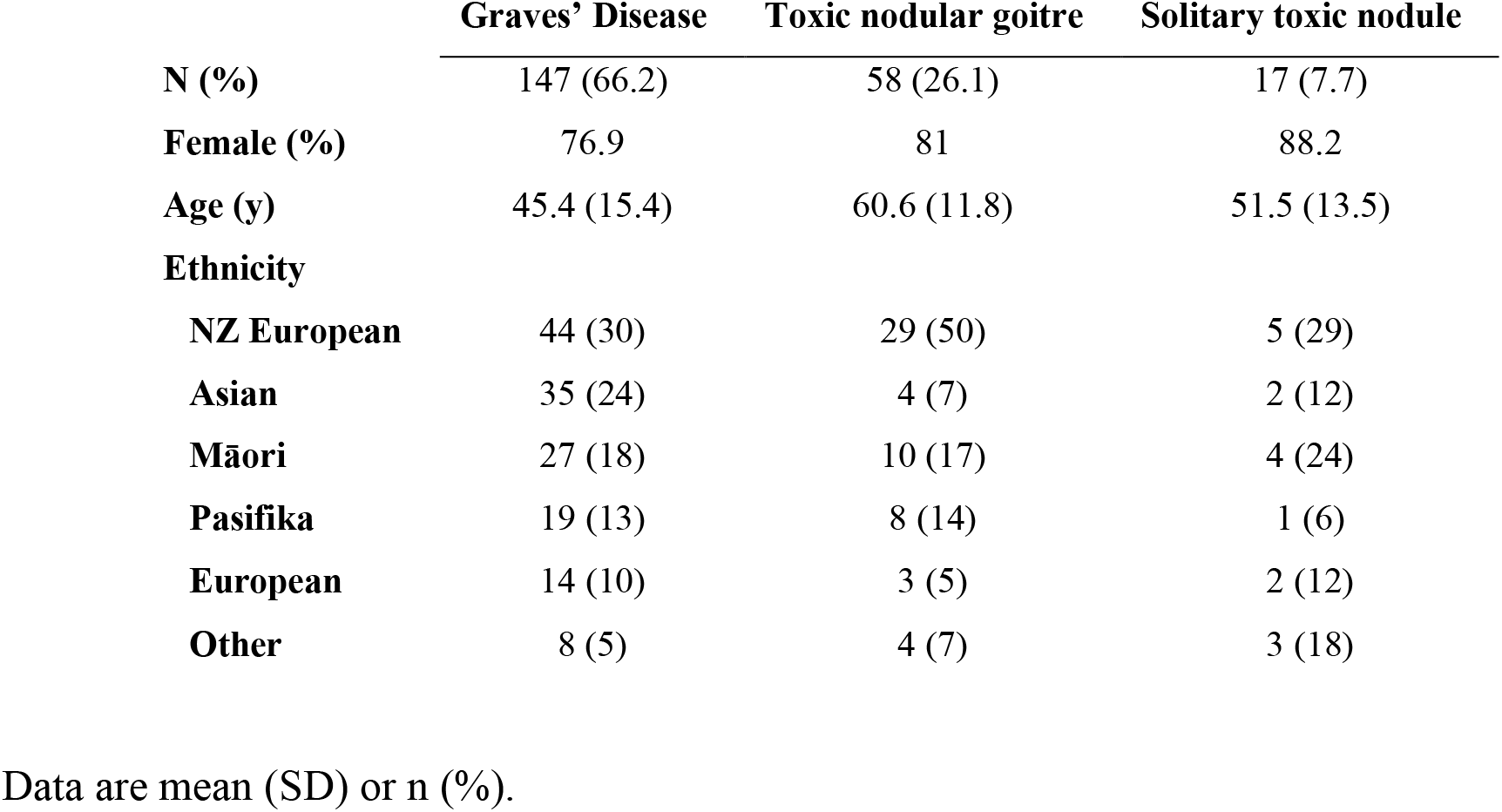
selected characteristics of the cohort prescribed radioiodine for thyrotoxicosis

|  | **Graves' Disease** | **Toxic nodular goitre** | **Solitary toxic nodule** |
| --- | --- | --- | --- |
| **N (%)** | 147 (66.2) | 58 (26.1) | 17 (7.7) |
| **Female (%)** | 76.9 | 81 | 88.2 |
| **Age (y)** | 45.4 (15.4) | 60.6 (11.8) | 51.5 (13.5) |
| **Ethnicity** |  |  |  |
| **NZ European** | 44 (30) | 29 (50) | 5 (29) |
| **Asian** | 35 (24) | 4 (7) | 2 (12) |
| **Māori** | 27 (18) | 10 (17) | 4 (24) |
| **Pasifika** | 19 (13) | 8 (14) | 1 (6) |
| **European** | 14 (10) | 3 (5) | 2 (12) |
| **Other** | 8 (5) | 4 (7) | 3 (18) |

Data are mean (SD) or n (%).

### Graves’ Disease

147 patients with Graves’ disease received radioiodine. Table 2 shows selected patient characteristics, treatment and outcomes. The average duration since diagnosis was 3.7 years; 103 (70%) had a measurement of TSH-R ab an average of 1.2 years before treatment; and for most patients (71%), the most recent TSH prior to radioiodine (taken an average of 33 days pre-treatment) was either undetectable or below the normal range. 90% of patients were taking anti-thyroid drugs with the median dose of carbimazole 15mg/day and propylthiouracil 250 mg/day.

**Table 2:** characteristics, treatment, and outcomes of the cohort by diagnosis

| **Characteristics** | **Graves' Disease**<br>**(n=147)** | **Toxic nodular**<br>**goitre (n=58)** | **Solitary toxic**<br>**nodule (n=17)** |
| --- | --- | --- | --- |
| Time since diagnosis (y)<sup>a</sup> | 3.7 (3.5) [0-21] | - | - |
| TSH receptor antibody (n=103, IU/L)<sup>b</sup> | 11.9 (10.6) | - | - |
| TSH prior to radioiodine<sup>c</sup> |  |  |  |
| Undetectable | 80 (54.4) | 24 (41.4) | 7 (41.2) |
| Suppressed (<0.4 mU/L) | 25 (17.0) | 21 (36.2) | 7 (41.2) |
| Normal (0.4-4 mU/L) | 28 (19.0) | 13 (22.4) | 3 (17.6) |
| High (>0.4 mU/L) | 14 (9.5) | 0 (0.0) | 0 (0.0) |
| Medication use |  |  |  |
| Carbimazole<sup>c</sup> | 118 (80.3) | 39 (67.2) | 9 (52.9) |
| Daily dose (mg/day)<sup>d</sup> | 15 [1.25-60] | 10 [2.5-60] | 10 [5-10] |
| Propylthiouracil<sup>c</sup> | 14 (9.5) | 2 (3.4) | 0 (0.0) |
| Daily dose (mg/day)<sup>d</sup> | 250 [100-450] | 125 [50-200] | - |
| Nil<sup>c</sup> | 15 (10.2) | 17 (29.3) | 8 (47.1) |
| Radioiodine treatment<sup>c</sup> |  |  |  |
| One dose | 118 (80.3) | 54 (93.1) | 17 (100) |
| Two doses | 27 (18.4) | 4 (6.9) | 0 (0.0) |
| Three doses | 2 (1.4) | 0 (0.0) | 0 (0.0) |
| First dose (MBq)<sup>d</sup> | 550 [200-1000] | 550 [400-1000] | 550 [500-550] |
| Total dose (MBq)<sup>d</sup> | 550 [200-2400] | 550 [400-1800] | 550 [500-550] |
| Cure of Thyrotoxicosis<sup>c</sup> | 135 (91.8) | 54 (93.1) | 17 (100) |
| Required thyroxine after radioiodine<sup>c</sup> | 122 (83.0) | 13 (22.4) | 6 (35.3) |
| Elevated final TSH<sup>c</sup> | 51 (34.7) | 10 (17.2) | 8 (47.1) |

<sup>a</sup>Data are mean (SD) [range]; <sup>b</sup>mean (SD); <sup>c</sup>n (%); <sup>d</sup>median [range]

Most (80%) patients had 1 dose of radioiodine. The median first dose was 550 MBq, ranging from 200-1000 MBq. 27 patients had 2 doses, and two had 3 doses of radioiodine (median duration between doses 336 days, range 184-1098 days). The range of total dose received was 200-2400 MBq. Overall, 92% of patients had the thyrotoxicosis cured, and 83% required thyroxine post-radioiodine. All but one patient had a TSH measurement after radioiodine, on average the last measurement 3.2 years after their last radioiodine dose. In 51 (35%) people, this final TSH measurement was above the normal range. Only 13 patients (9%) were considered cured after treatment and did not require thyroxine treatment and had a normal final TSH measurement.

Table 3 shows treatment and outcome data by the first dose of radioiodine received. The most commonly prescribed doses were 400, 500, 550, and 600 MBq. Generally, the cure rates for thyrotoxicosis, and requirement for thyroxine decreased with increasing first doses. Of those receiving ≤500 MBq for the first dose, the cure rate was >80% after the first dose, about 75% required thyroxine after the first dose, 9 (12.5%) required a second dose, and 8 of these 9 patients were cured. Of those receiving ≥550 MBq for the first dose, the initial cure rates were generally <70%, and overall >25% required a second dose. Neither age, gender, nor ethnicity were associated with likelihood of cure, but there were only a small proportion of individuals who were not cured meaning these results should be interpreted cautiously.

**Table 3:**
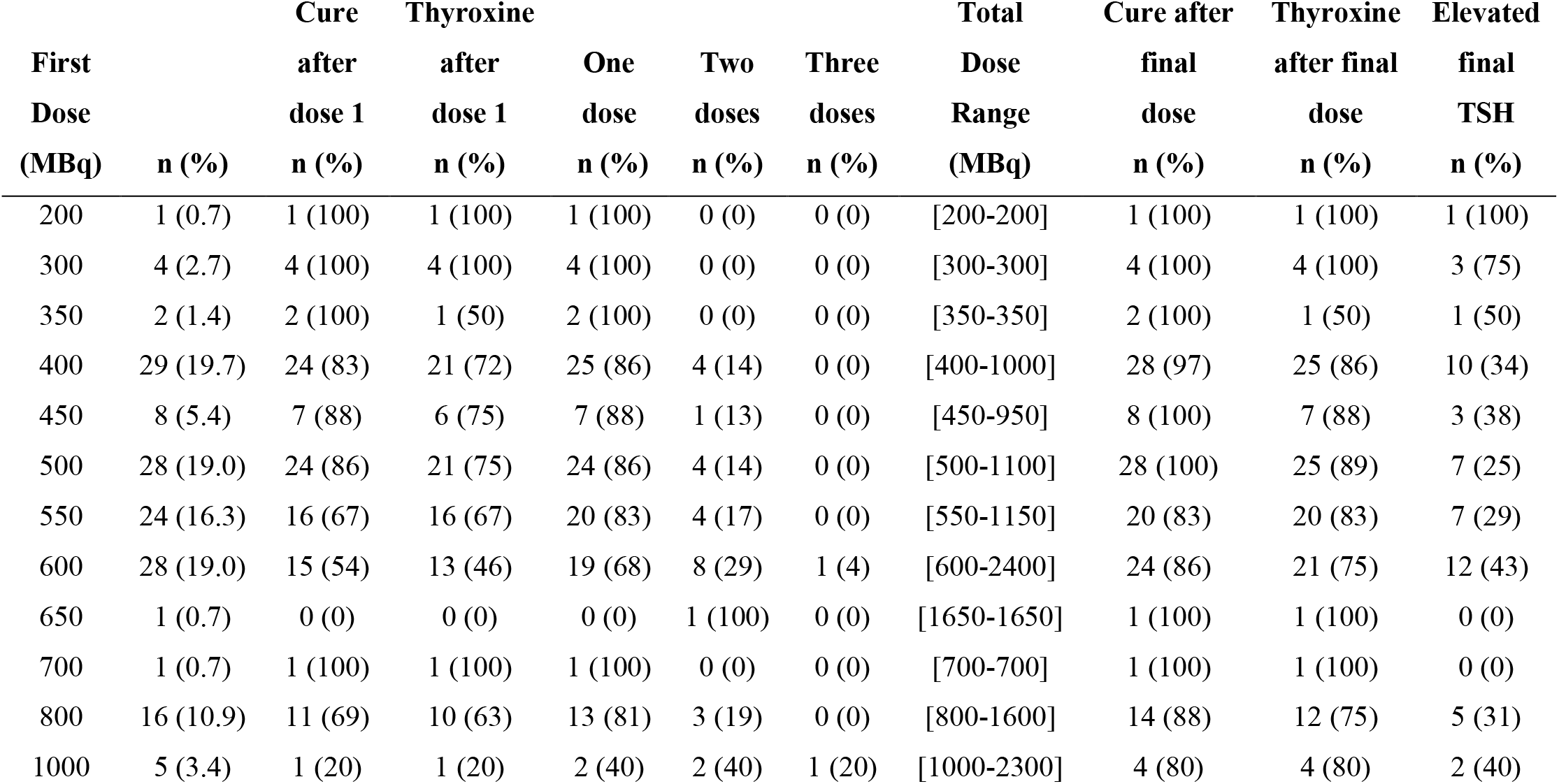
treatment and outcomes in Graves’ disease by initial radioiodine dose

We assessed potential factors that might influence the development of hypothyroidism after radioiodine. Pre-radioiodine treatment with carbimazole or propylthiouracil in doses of at least 15 mg/day or 150 mg/day respectively was not associated with incidence of hypothyroidism (83% on high dose medication vs 84% on low dose/no medication, P>0.99). Pre-radioiodine TSH predicted development of hypothyroidism (TSH undetectable or suppressed, ie <0.4 mU/L, 78% vs TSH normal or elevated 95%, P=0.006). Higher radioiodine total dose was associated with numerically higher rates of hypothyroidism development, but the differences were not statistically significant. Using a threshold of >800 MBq, 30 patients received a total dose ≥900 MBq (2 patients one dose of 1000 MBq, remainder multiple doses) and 28 (93%) developed hypothyroidism vs 94/117 (80%) of patients receiving ≤800 MBq (1 patient received two 400 MBq doses, remainder received one dose), P=0.06.

### Toxic nodular goitre/solitary toxic nodule

58 patients with toxic nodular goitre and 17 with a solitary toxic nodule received radioiodine. Table 2 shows selected patient characteristics, treatment and outcomes. Most patients (78-82%) had a pre-radioiodine TSH (average of 37-45 days pre-treatment) that was either undetectable or below the normal range. 29-47% of patients were not taking antithyroid drugs at the time of treatment.

Most (93%) patients with toxic nodular goitre and all patients with a solitary toxic nodule had only 1 dose of radioiodine. The median first dose was 550 MBq, ranging from 400-1000 MBq for toxic nodular goitre and 500-550 MBq for solitary toxic nodule. 4 patients with toxic nodular goitre had 2 doses, receiving a total dose of 1100-1800 MBq (median duration between doses 245 days, range 174-462 days). Overall, 93-100% of patients had the thyrotoxicosis cured. For toxic nodular goitre 22% required thyroxine post-radioiodine, and for solitary toxic nodule, 35% required thyroxine. A TSH above the normal range at the last measurement (on average 3.0-3.1 years after radioiodine) was present in 17% of those with toxic nodular goitre, and 47% with a solitary toxic nodule. 62% of those with toxic nodular goitre were considered cured after treatment and did not require thyroxine treatment and had a normal final TSH measurement, but only 29% of those with a solitary toxic nodule met those criteria.

Table 4 shows treatment and outcome data by the first dose of radioiodine received. The most commonly prescribed dose was 550 MBq. Generally, there were too few data otherwise to draw conclusions regarding dose and outcomes. We again assessed potential factors that might influence the development of hypothyroidism after radioiodine. None of pre-radioiodine medication dose, pre-radioiodine TSH, or radioiodine dose predicted hypothyroidism development, but these analyses should again be interpreted cautiously because of the small number of cases in the various groups.

**Table 4:**
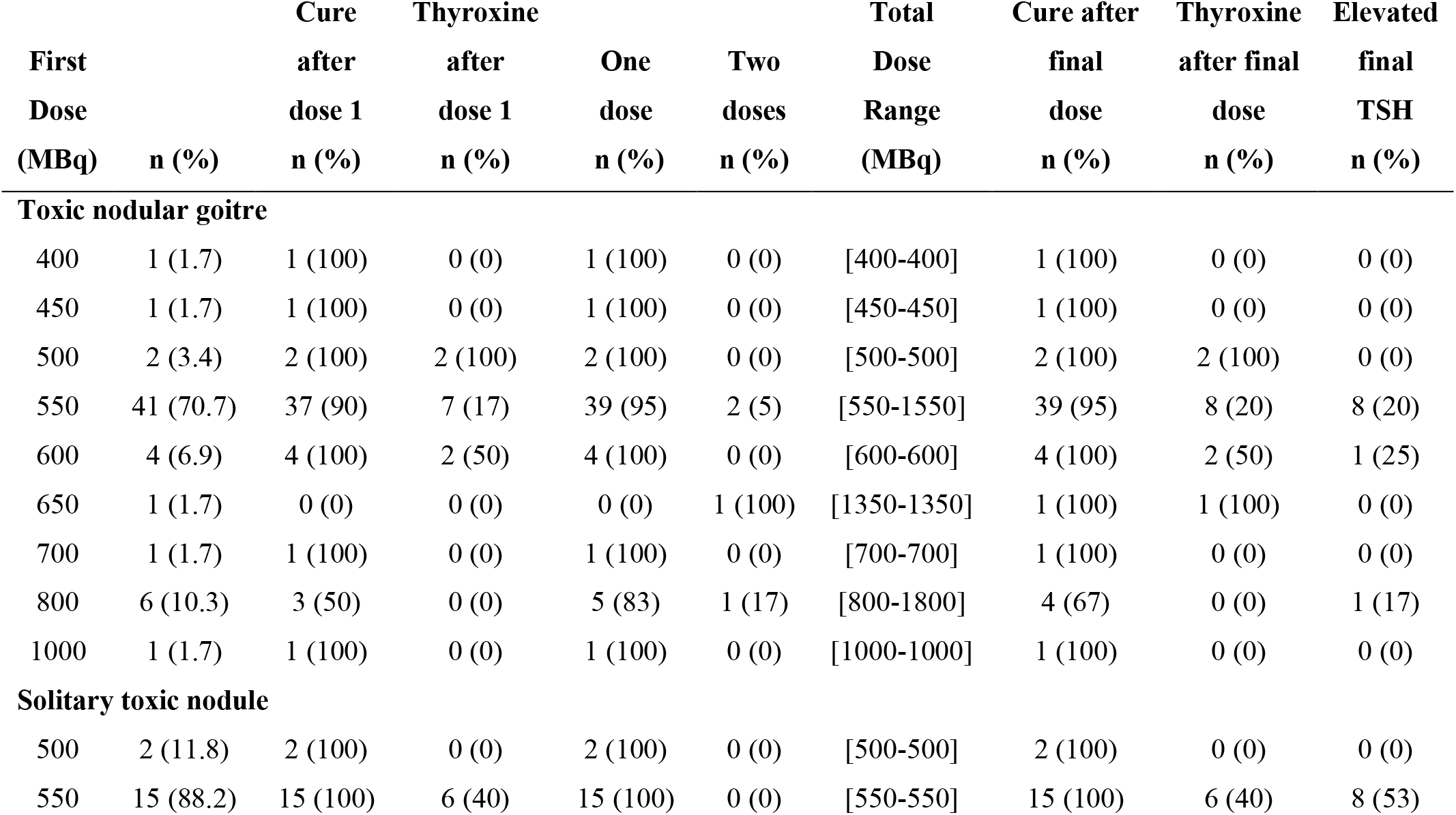
treatment and outcomes in toxic nodular goitre and solitary toxic nodule by initial radioiodine dose

| First Dose (MBq) | n (%) | Cure after dose 1 n (%) | Thyroxine after dose 1 n (%) | One dose n (%) | Two doses n (%) | Total Dose Range (MBq) | Cure after final dose n (%) | Thyroxine after final dose n (%) | Elevated final TSH n (%) |
| --- | --- | --- | --- | --- | --- | --- | --- | --- | --- |
| **Toxic nodular goitre** |  |  |  |  |  |  |  |  |  |
| 400 | 1 (1.7) | 1 (100) | 0 (0) | 1 (100) | 0 (0) | [400-400] | 1 (100) | 0 (0) | 0 (0) |
| 450 | 1 (1.7) | 1 (100) | 0 (0) | 1 (100) | 0 (0) | [450-450] | 1 (100) | 0 (0) | 0 (0) |
| 500 | 2 (3.4) | 2 (100) | 2 (100) | 2 (100) | 0 (0) | [500-500] | 2 (100) | 2 (100) | 0 (0) |
| 550 | 41 (70.7) | 37 (90) | 7 (17) | 39 (95) | 2 (5) | [550-1550] | 39 (95) | 8 (20) | 8 (20) |
| 600 | 4 (6.9) | 4 (100) | 2 (50) | 4 (100) | 0 (0) | [600-600] | 4 (100) | 2 (50) | 1 (25) |
| 650 | 1 (1.7) | 0 (0) | 0 (0) | 0 (0) | 1 (100) | [1350-1350] | 1 (100) | 1 (100) | 0 (0) |
| 700 | 1 (1.7) | 1 (100) | 0 (0) | 1 (100) | 0 (0) | [700-700] | 1 (100) | 0 (0) | 0 (0) |
| 800 | 6 (10.3) | 3 (50) | 0 (0) | 5 (83) | 1 (17) | [800-1800] | 4 (67) | 0 (0) | 1 (17) |
| 1000 | 1 (1.7) | 1 (100) | 0 (0) | 1 (100) | 0 (0) | [1000-1000] | 1 (100) | 0 (0) | 0 (0) |
| **Solitary toxic nodule** |  |  |  |  |  |  |  |  |  |
| 500 | 2 (11.8) | 2 (100) | 0 (0) | 2 (100) | 0 (0) | [500-500] | 2 (100) | 0 (0) | 0 (0) |
| 550 | 15 (88.2) | 15 (100) | 6 (40) | 15 (100) | 0 (0) | [550-550] | 15 (100) | 6 (40) | 8 (53) |

### Hypothyroidism at the final visit

69 (31%) of patients had an elevated TSH at the final visit (35%, 17%, 47% for Graves’ disease, toxic nodular goitre, and solitary toxic nodule respectively). Of these 69 patients, in 21 (30%), the TSH was >10 mU/L, and in 48 (70%) it was between 4-10 mU/L. 52/69 (75%) were taking thyroxine (19/21 with TSH >10 mU/L, and 33/48 with TSH 4-10 mU/L), and overall, 52/141 (37%) patients taking thyroxine had a final elevated TSH. Neither age, gender nor ethnicity were associated with the presence of hypothyroidism at the final visit.

In patients with Graves’ disease and elevated TSH, 19 had TSH >10 mU/L (17 on thyroxine) and 32 had TSH 4-10 mU/L (28 on thyroxine). For toxic nodular goitre, 1 had TSH >10 mU/L (on thyroxine) and 9 had TSH 4-10 mU/L (4 on thyroxine), and for solitary toxic nodule, 1 had TSH >10 mU/L (on thyroxine), and 7 had TSH 4-10 mU/L (1 on thyroxine).

## Discussion

In this review of 222 patients with thyrotoxicosis receiving radioiodine at ADHB in 2015, more than 90% were ultimately cured. In Graves’ disease, multiple doses of radioiodine were required in 20%, whereas in toxic nodular goitre (7%) and solitary toxic nodule (0%) the proportions receiving multiple doses were lower. In Graves’ disease, most individuals (83%) required thyroxine after radioiodine, whereas in toxic nodular goitre (22%) and solitary toxic nodule (35%) the proportions requiring thyroxine were lower but not insignificant. A substantial minority of patients (31%) had elevated TSH levels at their most recent blood test, and in 30% of these individuals, the TSH was >10 mU/L. Most patients with elevated TSH levels (75%) were taking thyroxine, and, overall, 37% of patients taking thyroxine had a final elevated TSH. In Graves’ disease, a normal or elevated pre-radioiodine TSH or a higher total dose received tended to be associated with higher rates of post-radioiodine hypothyroidism, but the increases in rates were only small. Overall, the chance of being cured of thyrotoxicosis without need for thyroxine and with a normal TSH was <10% in Graves’ disease, <30% in solitary toxic nodule, but much higher (62%) in toxic nodular goitre. These rates of achieved euthyroidism are considerably lower than physician expectations in the Australian survey of endocrinologists, in which 40% aimed for euthyroidism after radioiodine for Graves’ disease.^1^

We identified four reports of outcomes following radioiodine treatment for thyrotoxicosis in Australia or New Zealand. Gupta and colleagues reported rates of cure and euthyroidism at 4 months in 478 patients with thyrotoxicosis treated in Newcastle with a single fixed dose of 555 MBq.^3^ Overall rates of cure and hypothyroidism were 79.5% and 35.4% respectively (61%, 46% Graves’; 87%, 30% toxic nodular goitre; 90%, 18% solitary toxic nodule). Notably <25% of the cohort had Graves’ disease (compared to 66% in our cohort). Fanning and colleagues reported similar data for 92 patients with Graves’ disease treated in Brisbane with a fixed dose of 450-500 MBq and followed for at least 12 months.^4^ 79% were cured after one dose (70% hypothyroid) and 12 patients had a second dose, all of whom were cured and developed hypothyroidism (a further 2 patients were cured with surgery after their first dose). Overall, 85/90 (94%) were cured with radioiodine alone, but 81/90 (90%) developed hypothyroidism. Finally, Tamatea and colleagues reported outcome data for 223 patients with thyrotoxicosis between 2008-10 followed for a median of 2.5 years and 589 patients with Graves’ disease between 2001-13 followed for up to 10 years treated in Hamilton, New Zealand, with a fixed dose of 555 MBq.^5,6^ 78.5% were cured after one dose (76% Graves’ disease, 82% toxic nodular goitre/solitary toxic nodule), but cure was substantially lower in Māori.^5^ In prolonged follow-up the proportion of patients with TSH >5 mU/L remained between 25-30% at 1, 2, 5, and 10 years after treatment, and 41-61% of those with elevated TSH had a TSH >10 mU/L.^6^

At ADHB, individualised rather than fixed doses of radioiodine were prescribed. Therefore, relationships between dose prescribed and outcomes are difficult to interpret because they are affected by indication bias. Individuals receiving higher doses were prescribed those doses because, based on the patient characteristics^8^ (eg size of goitre, presence of elevated TSH-R ab etc), the clinician thought it less likely that cure would occur with a lower dose. It is therefore unsurprising that cure rates and need for thyroxine after the first dose was lower in patients with Graves’ disease receiving higher doses of radioiodine.

Few randomised controlled outcome data are available to guide clinicians about radioiodine dosing. Although patient characteristics such as the thyroid size predict outcomes following radioiodine,^3^ randomised trials of radioiodine doses calculated based on thyroid size or uptake of iodine do not show better outcomes than those using a fixed dose of radioiodine.^9,10^ For fixed doses, randomised trials have reported higher cures rates for single doses of 370 vs 185 MBq (88.5 vs 48.5% at 2 years),^11^ and for 1110 vs 555 MBq (94.8% vs 77.4% at 1 year),^12^ but no difference between 235 vs 350 MBq (73% vs 74% after mean 63 months),^9^ or between 370 vs 555 MBq (73.7% vs 80% after 19 months).^13^

Ideally, prescribing would be guided by evidence derived from randomised trials, but it seems unlikely that adequately powered trials comparing outcomes between small differences in doses would be conducted because the likely differences in outcomes between such doses would be small. Therefore, individualised dosing seems likely to continue to be somewhat pragmatic, but it seems reasonable to rationalise it to standard fixed doses eg 200, 400, 550 or 600, 800, 1000 MBq, depending on thyroid size and other patient characteristics. Using doses outside of such standard doses seems unlikely to provide clinically relevant differences in cure rates or need for thyroxine.

Distancing and hygiene precautions are required after radioiodine. In our practice, doses ≤400 MBq are required to maintain strict 2m distance precautions for 2 days, doses 400-600 MBq for 4 days and >600 MBq for 6-8 days or longer. Individualised dosing means that individuals prescribed smaller doses of radioiodine will benefit from shorter isolation periods, but conversely those prescribed higher doses are required to have longer periods.

31% of patients overall and 37% taking thyroxine had elevated TSH levels at their final measurement, and about 30% were >10 mU/L. These findings mirror the results of the audit from Hamilton New Zealand, which also found a substantial proportion of patients with elevated TSH levels after radioiodine treatment.^6^ Mildly elevated TSH levels are unlikely to cause symptoms or affect health, but in both audits a substantial number of patients had TSH >10 mU/L which may impact on health. Given the likelihood of long-term thyroxine use was very high in Graves’ disease (83%) and not uncommon in toxic nodular goitre (22%) and solitary toxic nodule (35%), it is essential that patients be well informed about this possibility during discussions about radioiodine treatment, and that physicians are realistic about the likelihood of requiring thyroxine. The Australian survey of endocrinologists’ practice in which 40% aimed for euthyroidism^1^ suggests that many physicians underestimate this possibility. As many patients are already taking multiple tablets of antithyroid medication per day or are also considering thyroidectomy after which thyroxine will universally be required, the point may seem moot because taking and monitoring thyroxine is probably easier than taking and monitoring antithyroid drugs. However, many patients hope that they will be medication-free with normal thyroid function after radioiodine. The current data show that this is unlikely (<10%) in Graves’ disease, surprisingly uncommon in solitary toxic nodule (<30%) and only occurs in about 2/3 with toxic nodular goitre.

## Conclusion

This audit provides outcome data on individualised treatment of radioiodine for thyrotoxicosis. Taken together with the previous published data on fixed doses from Australia and New Zealand, they provide a reference for cure rates and requirement for thyroxine after radioiodine for Graves’ disease, toxic nodular goitre and solitary toxic nodule. The data also suggest that a substantial proportion of individuals taking thyroxine have suboptimal control of their hypothyroidism.

## Data Availability

Data is available from the corresponding author on reasonable request.

## Notes

**Competing interests:** All authors have completed the ICMJE uniform disclosure form at www.icmje.org/coi_disclosure.pdf and declare: no support from any organization for the submitted work; no financial relationships with any organizations that might have an interest in the submitted work in the previous three years; no other relationships or activities that could appear to have influenced the submitted work.”

### Competing Interest Statement

The authors have declared no competing interest.

### Funding Statement

The study received no specific funding but MJB is the recipient of an HRC Clinical Practitioner Fellowship.

### Author Declarations

This is an audit as defined by the New Zealand National Ethics Advisory Committee guidelines and therefore it did not require ethical approval. As such, ethical approval was waived by the Health and Disability Ethics Committees (HDEC) of New Zealand.

